# Evaluating a communication aid for return of genetic results in families with hypertrophic cardiomyopathy: A randomized controlled trial

**DOI:** 10.1101/2021.12.19.21268076

**Authors:** Charlotte Burns, Laura Yeates, Joanna Sweeting, Christopher Semsarian, Jodie Ingles

**Author notes:** **Corresponding author:** A/Prof Jodie Ingles, Centre for Population Genomics, Garvan Institute of Medical Research, 384 Victoria St, Darlinghurst, NSW 2010, Twitter: @jodieingles27. **Ethics:** Participants provided written informed consent. Ethical approval was granted by the Sydney Local Health District Human Research Ethics Review Committee (X16-0030). **Trial registration:** The trial was registered with the Australian New Zealand Clinical Trials Registry: ACTRN12617000706370.

## Abstract

**Introduction:** Genetic testing for hypertrophic cardiomyopathy (HCM) is considered a key aspect of management. Communication of genetic test results to the proband and their family members, can be a barrier to effective uptake. We hypothesized that a communication aid would facilitate effective communication, and sought to evaluate knowledge and communication of HCM risk to at-risk relatives.

**Methods:** This was a prospective randomized controlled trial. Consecutive HCM patients attending a specialized clinic, who agreed to participate, were randomized to the intervention or current clinical practice. The intervention consisted of a genetic counselor-led appointment, separate to their clinical cardiology review, and guided by a communication booklet which could be written in and taken home. Current clinical practice was defined as the return of the genetic result by a genetic counselor and cardiologist, often as part of a clinical cardiology review. The primary outcome was the ability and confidence of the individual to communicate genetic results to at-risk relatives.

**Results:** The *a priori* outcome of improved communication amongst HCM families did not show statistically significant differences between the control and intervention group, though the majority of probands in the intervention group achieved fair communication (n=13/22) and had higher genetic knowledge scores than those in the control group (7 ± 3 versus 6 ± 3). A total of 29% of at-risk relatives were not informed of a genetic result in their family.

**Conclusion:** Communication amongst HCM families remains challenging, with nearly a third of at-risk relatives not informed of a genetic result. We show a significant gap in the current approach to supporting family communication about genetics.

Australian New Zealand Clinical Trials Registry: ACTRN12617000706370

**What is known about this topic:** Communication between family members regarding genetic testing can be a barrier to effective uptake.

**What this paper adds to the topic:** Communication is a challenge in HCM families, with nearly a third of at-risk relatives not informed of a genetic result. There is a significant gap in the current approaches to supporting family communication about genetics.

## INTRODUCTION

Hypertrophic cardiomyopathy (HCM) is a clinically heterogeneous disease characterised by unexplained left ventricular hypertrophy in the absence of a loading condition such as hypertension (Maron et al., 2012). HCM affects at least 1 in 500 with some being relatively asymptomatic, while others can have heart failure or sudden cardiac death (Ho et al., 2018; Semsarian et al., 2015). HCM genetic testing can allow identification of a causative genetic variant, which can be used for cascade genetic testing and clarifying risk status of family members (Ingles, Bagnall, et al., 2018; Ingles et al., 2015). Effective pre-test and post-test genetic counseling is critical and recognised in disease guidelines, and focuses on clear communication of the likely outcomes of HCM genetic testing (Burns et al., 2018).

Genetic counseling is a critical aspect of the process, not just for genetic testing and explaining uncertain and complex results, but also for understanding inheritance risks, characterisation of the family history, information and emotional support (Burns et al., 2018; Ingles & Semsarian, 2018). In the clinic setting, pre- and post-test genetic counseling should include discussion of inheritance risks and clinical screening guidelines for at-risk relatives (Burns et al., 2018; Semsarian et al., 2017). This allows asymptomatic at-risk relatives to make proactive, informed decisions regarding their risk of developing disease, including family planning decisions (Ingles & Semsarian, 2018). How a patient understands and communicates this genetic information to their at-risk relatives is critical to ensuring patients’ get the most value out of genetic testing. Communication with the wider family relies on the proband and several studies highlight the many ways this can be problematic, including that individuals may not retain or understand the information presented to them (Burns et al., 2018; Burns et al., 2016; Burns, Yeates, et al., 2017; Patenaude et al., 2013; Young et al., 2017).

It is estimated 20-40% of relatives are unaware of relevant genetic information and do not act on information even when they have reportedly been informed of their risk (Burns et al., 2016; Christiaans et al., 2008; Gaff et al., 2007). In a qualitative study of HCM patients undergoing comprehensive genetic testing, many patients report uncertain results to be conveyed less amongst families (Burns, Yeates, et al., 2017). The general genetics literature highlights that risk perception and understanding of results though varied, can be poor, inaccurate and incomplete (Patenaude et al., 2013; Young et al., 2017).

Decision or communication aids are tools specifically designed to support patients with decision making and unmet information needs. They have been shown to be effective in improving knowledge and accuracy of risk perceptions (Stacey et al., 2017; Vavolizza et al., 2015; Wakefield et al., 2007). Few resources exist that aim to facilitate effective communication to relatives at risk of HCM. We hypothesize that improving knowledge of an HCM genetic result would have a positive impact on communication to at-risk relatives. We aim to evaluate knowledge and communication of HCM risk to relatives following use of a custom designed HCM decision aid. The decision aid has been previously developed and trialed to demonstrate feasibility and acceptability (Smagarinsky et al., 2017).

## METHODS

### Study design

This was a prospective randomized controlled trial and the protocol has been previously published (Burns et al., 2019). The trial was registered with the Australian New Zealand Clinical Trials Registry: ACTRN12617000706370. Consecutive HCM patients were invited to participate when notified during their genetic counseling intake call that their genetic result was ready. Once verbal consent was obtained they were randomized to receive their genetic result via the intervention or control arm of the study (Figure 1). The local Human Research Ethics Committee approved the study (Sydney Local Health District Ethics Review Committee; X16-0030).

**Figure 1.**
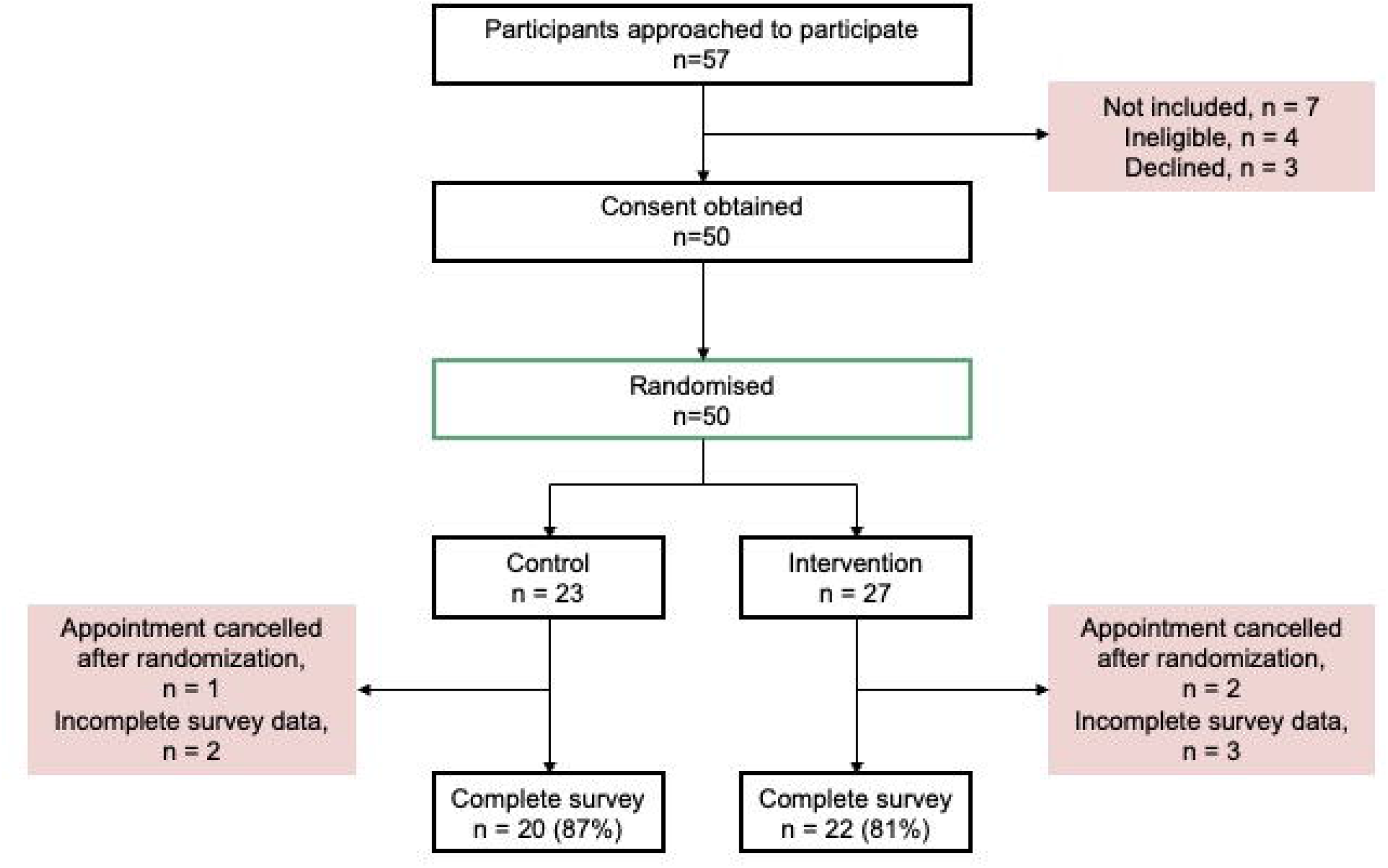
Flowchart of Randomized Controlled Trial Study Probands.

### Participants

This trial was conducted in a specialized multidisciplinary HCM clinic at the Royal Prince Alfred Hospital, Sydney, Australia. HCM probands with a genetic result ready for return were eligible. Participants were aged 18 years or older, with sufficient written English skills. Genetic testing was performed as part of a research study, or by a clinically accredited laboratory, as previously described (Bagnall et al., 2016; Burns, Bagnall, et al., 2017). Variants were classified using the *MYH7*-modified ACMG/AMP criteria (Bagnall et al., 2016; Burns, Bagnall, et al., 2017). Recruitment commenced in November 2017 and ended in November 2018. Invitation to participate occurred during the genetic counseling pre-clinic intake phone call conducted as per regular clinical process, and participants indicated verbally if they were interested in participating. Informed written consent was obtained by the cardiac genetic counselor present at the participants’ clinic consultation.

### Randomization

A randomized list was prepared using the Excel (Microsoft Office) “Random” function and study participants were allocated the next number on the random list. This number was linked to either control or intervention. A researcher not involved in the study performed the randomization to ensure allocation concealment.

### Sample Size and Power Calculations

Prior to commencement of the study, sample size calculations were performed using the results from our published feasibility study (Smagarinsky et al., 2017). The primary outcome of this trial was the ability and confidence of the proband to communicate genetic results to at-risk relatives. Data from the feasibility study indicated 75% of participants communicated genetic results to at-risk relatives. Assuming the control group communicates in 50% of cases, at a significance level of 5% and 80% statistical power, a sample size of n=21 was required per group. We planned to recruit additional patients with the aim of reducing the impact of drop-out and incomplete survey data on endpoints.

### Development of the Communication Aid

We previously developed a communication aid to assist with the delivery of genetic results to the proband and to support family communication. The communication aid published and reported in the pilot study by Smagarinsky et al was used in this current randomized controlled trial (Smagarinsky et al., 2017). The pilot data demonstrated feasibility and acceptability of the communication aid for use in such a trial.

### Control Group

Those in the control group received their result as per current clinical practice. In our practice, this typically involves return of a genetic result either by the cardiologist or genetic counselor. Return of the result is usually performed following clinical cardiology review, which is often the primary purpose of the consult. In the majority of cases a genetic counselor is present.

### Intervention Group

Those randomized to the intervention were allocated a separate appointment time after clinical review with their cardiologist. In this appointment they saw the cardiac genetic counselor who returned their genetic result using the communication aid.

The communication aid covers the process of genetic testing through to the implications of a genetic result for at-risk relatives (Burns et al., 2019). There is a section in the aid under ‘Results’, which goes through the meaning of each category of genetic result. These include an indeterminate result (no variant identified), a variant of uncertain significance and a likely pathogenic/pathogenic result. The genetic counselor returning the genetic result marked the appropriate category of result and specific recommendations for the rest of the family.

### Data Collection and Outcomes

Both the primary and secondary outcomes were measured at a single time point (two-weeks post intervention) using a survey comprising a number of previously published and validated scales. A number of demographic questions were asked within the survey. The survey was available online via Qualtrics (https://www.qualtrics.com/) with a direct link sent to participants. For those who preferred a hard copy it was posted with a return envelope. Evidence regarding the most appropriate time between genetic result disclosure and family communication is lacking. However, given the risk of arrhythmia and sudden death in the inherited heart disease context, two-weeks post result disclosure was considered by the study team to be an appropriate time point (Vavolizza et al., 2015). Return of the survey was followed up on a fortnightly basis.

### Primary Outcome

The primary outcome was the ability and confidence of the proband to communicate genetic results to at-risk relatives. This was measured at a single time point, and collected two-weeks after return of genetic results. Ability and confidence were assessed by two measures and then combined into a binary outcome. The certainty sub-scale of the Psychological Adaptation to Genetic Information (PAGIS) scale was used to measure confidence with genetic knowledge (Read et al., 2005). This sub-scale measures the patients’ perception and confidence in their genetic knowledge. Subsequent ability to pass this information on was measured by the percentage of at-risk relatives informed of genetic results by the proband. The percentage was calculated by counting the living first-degree relatives informed of their risk, and dividing by the total number of living first-degree relatives. We then averaged the scores from both measures to determine a final score. The final score was converted to a binary outcome of “fair” versus “poor” ability and confidence to communicate genetic results to at-risk relatives.

Fair communication was considered an average score of 75% and over, and poor communication <75%. We came to this cut-off after review of the literature and determined that communication rates fall between 60-80% but more often below 75%. In addition, we reviewed data from our previous studies in the field that showed similar rates of non-communication (Burns et al., 2016).

Factors that influence communication of genetic results to at-risk relatives are multidimensional. For this reason, we chose a combination approach to more broadly reflect the communication process. Many studies rely on single and linear measures of communication such as contact by relatives with genetics departments or self-reported communication with at-risk relatives only. To overcome this, we aimed to incorporate the proband’s confidence regarding their knowledge of genetics alongside the action linked to this knowledge, i.e. consistency between the probands confidence with their genetic information in combination with their self-reported percentage of immediate family members informed.

The certainty sub-scale of the PAGIS was used to measure confidence with genetic knowledge as described above (Read et al., 2005). Guided by grounded theory in patient perspectives of genetic counseling and the Roy Adaptation to Genetic Information Model, the 26-item PAGIS scale allows for evaluation of the efficacy of genetic counselling (Kasparian et al., 2007; Read et al., 2005). The scale aims to incorporate the multidimensional adaptation to genetic information and comprises five domains which include; a) non-intrusiveness, b) support c) self-worth, d) certainty and e) self-efficacy (Read et al., 2005). Evidence for the utility of this scale has been published and illustrates its potential use for assessing genetic counseling interventions (Read et al., 2005).

### Secondary Outcomes

The survey consisted of three additional scales to assess secondary outcomes, a number of questions regarding communication with relatives, as well as a number of demographic questions.

Genetic knowledge was assessed using an amended version of the Breast Cancer Genetic Counseling Knowledge Questionnaire (BGKQ) (Erblich et al., 2005; Kasparian et al., 2007). This scale was originally developed to assess knowledge of information typically included in genetic counseling for breast cancer. The original scale was a 27-item questionnaire including statements regarding genetics such as ‘*50% (half) of your genetic information was passed down from your mother’* and participants were asked if the statement was true or false. Items in the original scale were empirically derived from detailed content analysis of breast cancer genetic counseling sessions. The original scale demonstrated a high content validity with Cronbach’s α = 0.92, with demonstrated ability to discriminate between patients before and after genetic counseling sessions (Erblich et al., 2005). We amended questions to reflect the HCM context and 10 items were included.

Satisfaction with services received was assessed using the widely utilised Satisfaction with Genetic Counseling Scale (SGCS) (Shiloh et al., 1990). The original survey was designed to assess three dimensions of patient satisfaction: instrumental, affective and procedural (Kasparian et al., 2007; Shiloh et al., 1990). We used an amended version of the 12-item short form of the survey. We amended the scale by reducing it to nine questions therefore removing the procedural dimension to the scale (three questions). The authors of the scale advocate a flexible approach to scoring and we felt this met the needs of our study best (Shiloh et al., 1990).

The genetic counseling outcome scale (GCOS-24) was used to assess patient reported outcomes of genetic counselling (McAllister et al., 2011). The survey was designed to be used pre- and post-genetic counseling, though we have used it in the post-counseling setting to compare the control and intervention groups. The authors of this scale used the construct of empowerment to summarise the patient derived benefits from genetic counseling, suggesting a high score is indicative of patients feeling empowered with the information received in a genetic counseling session (McAllister et al., 2011).

### Data Analysis

Data were analysed using Prism (version 7.0) and SPSS (Version 23.0). We compared the primary outcome as a binary measure between the intervention and control group. We used chi-square analyses using p<0.05 for statistical significance. For assessment of secondary outcomes, we were guided by published scoring protocols for the validated scales to score genetics knowledge, satisfaction with services and genetic counseling outcomes. Mean scores for each scale were compared between the intervention and control group, and comparisons between the control and intervention group were analysed using unpaired t-tests for continuous data and chi-square analysis for categorical data. Sub-group analysis was performed; specifically, we compared outcomes in the study groups stratified by the gene result [informative (pathogenic/likely pathogenic)] and uninformative (uncertain or indeterminate)]. We also stratified probands by the presence or absence of a family history of disease.

## RESULTS

### Cohort Characteristics

We approached 57 eligible HCM probands with a genetic result ready to be returned. This included informative results (pathogenic/likely pathogenic) and uninformative results (variant of uncertain significance and indeterminate). Of those 57 probands, four were deemed ineligible due to insufficient English language skills and three declined. Fifty probands provided verbal consent to be randomized to the study. After randomization, three probands cancelled their clinic appointment. An additional five probands had insufficient data in their surveys to be included in analysis. This included three probands from the intervention arm and two probands from the control arm of the study. In total there were 20 probands in the control arm and 22 in the intervention arm (Figure 1).

Clinical and demographic characteristics of the probands included in the study are documented in Table 1. There were no statistically significant differences between probands in the control versus the intervention arms. Fifty percent (50%) of probands in the intervention arm and 50% of probands in the control arm had pathogenic/likely pathogenic variants identified. These figures accurately reflect the clinical yield of HCM genetic testing from the literature (Ho et al., 2018). The percent of at-risk first-degree relatives informed of a genetic result in the family was 71% (range: 0-100). This indicates 29% of at-risk relatives were not informed of a genetic result. The mean percentage of total at-risk first-degree relatives informed of their family members’ diagnosis of HCM was 83% (range: 0-100), indicating 17% were not informed.

**Table 1.** Participant characteristics.

|  | **Intervention** | **Control** | **p-value** |
| --- | --- | --- | --- |
| n | 22 | 20 | - |
| Male gender (%) | 16 (73) | 18 (90) | 0.24 |
| Current age (years) | 52 ± 16 | 50 ± 15 | 0.61 |
| European ethnicity (%) | 19 (86) | 14 (70) | 0.27 |
| Comorbidities present (%) | 4 (18) | 6 (30) | 0.48 |
| Pathogenic/likely pathogenic variant (%) | 11(50) | 10 (50) | 1 |
| ICD in situ (%) | 6 (23) | 9 (45) | 0.23 |
| SCD event (%) | 2 (9) | 1 (5) | 1 |
| Family history of clinical disease | 10 (45) | 6 (30) | 0.30 |
| Family history of SCD | 3 (14) | 3 (15) | 1 |
| Number living first-degree relatives (> 18 yrs) | 4.5 ± 2 | 3.5 ± 2 | 0.10 |

Abbreviations: ICD, implantable cardioverter defibrillator; SCD, sudden cardiac death.

### Primary Outcome

The *a priori* primary outcome measure was an average score which incorporated the certainty sub-scale from PAGIS and the number of first-degree relatives informed of their genetic test result. This was a binary score. Though more than half of participants in the intervention group demonstrated *“fair”* communication (≥75%) there was no statistically significant difference between the two groups (intervention:13/22 [59%] versus control:10/20 [50%], p= 0.26) (Figure 2). In addition, we compared the mean primary outcome score as a continuous variable and found no significant differences between the control and intervention groups (72 ± 4 versus 73 ± 4 years, p= 0.88).

**Figure 2.**
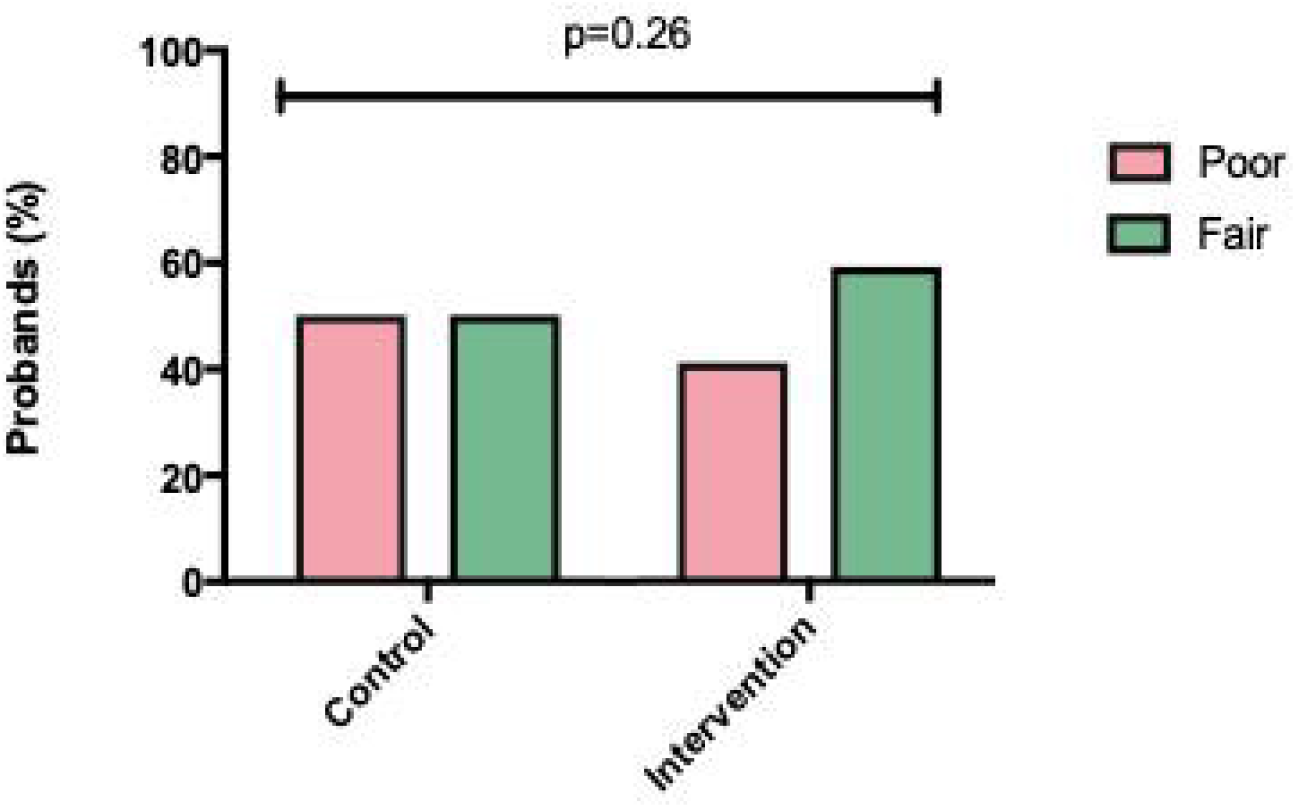
A priori primary outcome. Fair versus poor communication in control and intervention groups. The percentage of probands with fair versus poor communication in the control and intervention groups.

### Secondary Outcomes

#### Genetic Knowledge

The mean score for genetic knowledge amongst the total group was 6/10 (60%). Across the survey items, there were 107/420 ‘don’t know’ responses which were scored as incorrect (Figures 3A and B). There was no difference between probands in the intervention and control groups (Table 2). When considering the genetic knowledge score as a pass or fail, i.e. a score of less than 50% was considered a “fail” and >50% a “pass”, more of the intervention group received a pass for genetics knowledge though this did not reach statistical significance.

**Figure 3.**
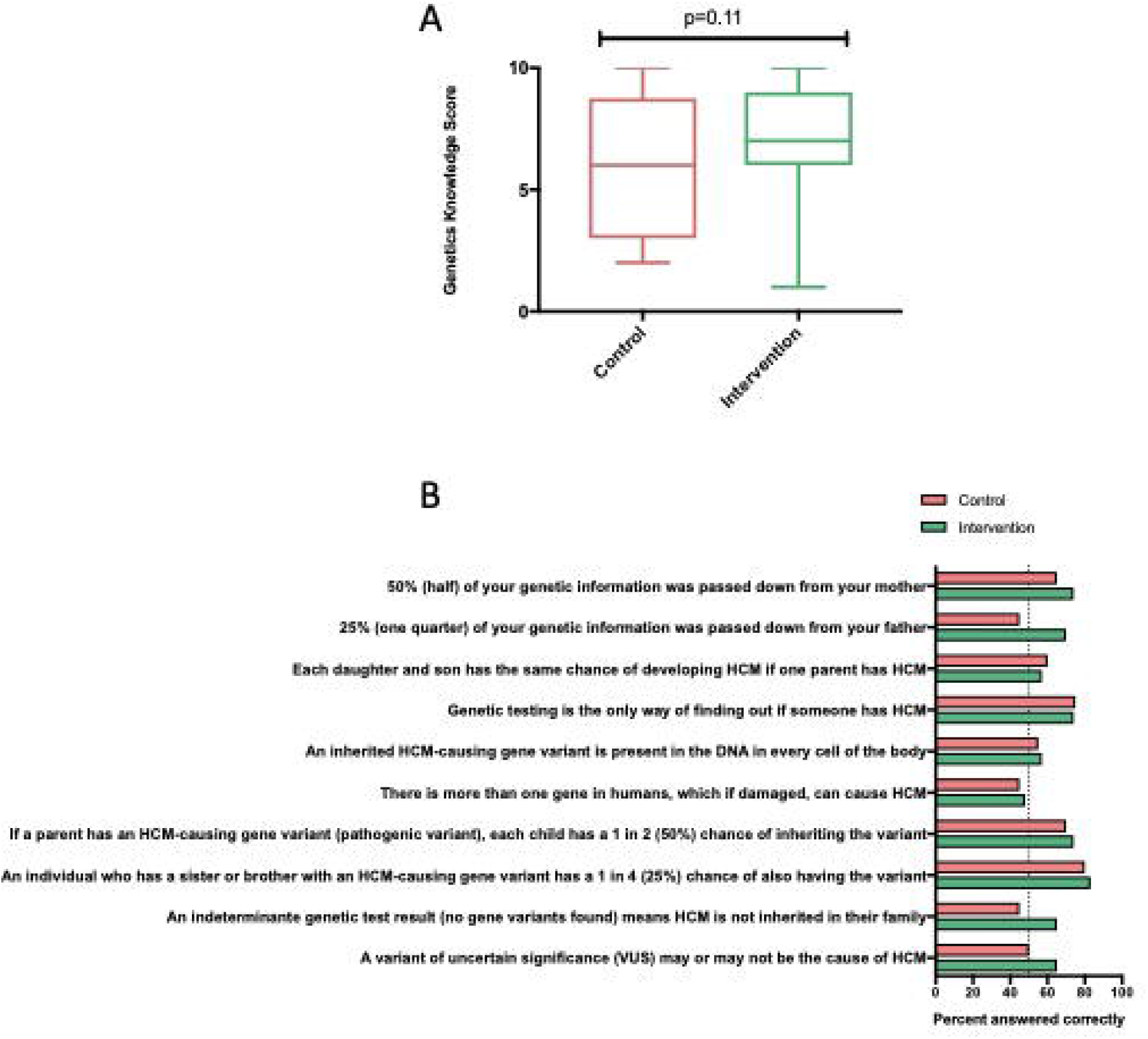
(A) t-test of genetic knowledge scores (maximum score of 10) between probands in the control and intervention groups. (B) Percentage of probands who gave correct answers for genetic knowledge items. Items from the genetic knowledge score answered correctly by control and intervention groups. Abbreviations: HCM = hypertrophic cardiomyopathy; DNA = deoxyribonucleic acid.

**Table 2.** Secondary Outcomes.

| Measure | Intervention | Control | p value |
| --- | --- | --- | --- |
| Genetic Knowledge (raw score) | 7 ± 3 | 6 ± 3 | 0.11 |
| Number with 'Pass' score (>50%) | 19/22 (86%) | 12/20 (60%) | 0.05 |
| PAGIS | 115 ± 16 | 112 ± 15 | 0.48 |
| Satisfaction with services | 33 ± 6 | 33 ± 4 | 0.91 |
| Instrumental | 11 ± 2 | 11 ± 1 | 0.90 |
| Affective | 11 ± 2 | 11 ± 1 | 0.76 |
| GCOS-24 | 120 ± 15 | 119 ± 16 | 0.72 |
| Understanding of need for referral | 6.7 ± 0.5 | 5.6 ± 1.0 | **0.01** |

Abbreviations: PAGIS, Psychological Adaptation to Genetic Information; GCOS-24, Genetic Counseling Outcomes Scale

#### Psychological Adaptation to Genetic Information

Overall, the mean total PAGIS score was 114 ± 16 (maximum score of 156) with higher total scores indicating more positive psychological adaptation amongst the group. When comparing the mean total score between the intervention and control groups there were no statistically significant differences (Table 2).

The mean total scores for the sub-scales included: non-intrusiveness 4.1 ± 0.8 (maximum weighted score of 6), support 4.5 ± 0.8 (maximum weighted score of 6), self-worth 4.4 ± 1.3 (maximum weighted score of 6) and self-efficacy 4.2 ± 1 (maximum weighted score of 6). The mean score for certainty was 4.4 ± 0.5 (maximum weighted score of 6), and was incorporated into the primary outcome calculation as described above. We compared the sub-scale scores between the intervention and control groups and there were no statistically significant differences.

#### Satisfaction with Services

Overall, all probands reported high levels of satisfaction with the process to return their genetic result. This was indicated by a mean total satisfaction score of 33 ± 5 (maximum score of 36). Higher scores indicate higher levels of satisfaction. For the instrumental and affective components of this scale, mean scores were 11 ± 2 (maximum score of 12) and 11 ± 2 (maximum score of 12) respectively. Single item scores (maximum score of 4) relating to expectations fulfilled, satisfaction with information and overall satisfaction all reflected high levels of satisfaction. When comparing the mean total score between the intervention and control groups there were no statistically significant differences (Table 2). We compared the instrumental and affective components between the intervention and control groups and there were no statistically significant differences (Table 2).

#### Patient Reported Outcomes of Genetic Counseling

The mean GCOS-24 score was 119 ± 15 (scores range from 24 to 168) which indicates good patient empowerment with higher scores indicating higher levels of empowerment (McAllister et al., 2011). When comparing the mean score between the intervention and control groups there were no statistically significant differences (Table 2). Probands in the intervention group were more likely to understand the reasons their doctor referred them to the cardiac genetic service (6.7 ± 0.5 versus 5.6 ± 1.0, p= 0.01) (Table 2).

## DISCUSSION

We describe a randomized controlled trial aimed at investigating the impact of a genetic counselor-led intervention to return HCM genetic results using a custom designed communication aid. The *a priori* primary outcome measure for this study was to assess the ability and confidence of the proband to communicate genetic results to at-risk relatives. Though this did not show statistical significance when compared between the intervention and control group, we highlight some important findings. First, the majority of participants in the intervention group did demonstrate *“fair”* communication as measured by the primary outcome and genetic knowledge scores were consistently higher amongst the intervention group. In addition, and of great clinical importance, we highlight that up to 29% of at-risk relatives remain uninformed about a genetic result in their family. Further, up to 17% of at-risk relatives remain uninformed of the HCM diagnosis itself. This is in spite of the return of results in a specialized multidisciplinary clinic with expertise including experienced cardiac genetic counselors and cardiologists. Uninformed relatives are unable to make proactive decisions regarding their own risk management. Factors influencing communication are multifaceted and likely require more than a single intervention to affect any meaningful improvement.

When asked about family communication, most patients report families should communicate risk amongst themselves with varying levels of support from their healthcare providers (K. Forrest et al., 2003; Healey et al., 2017; Young et al., 2017). In addition, there is evidence for the effectiveness of genetic counseling to assist with this process (Fiallos et al., 2017; L. E. Forrest et al., 2008; Healey et al., 2017). In spite of this, the literature consistently demonstrates family communication about genetics falls somewhere between 60-80% with a significant number of at-risk relatives remaining uninformed about their genetic risk (Gaff et al., 2007; Healey et al., 2017). At present, the biggest benefit of an informative genetic result is the opportunity for clinical and genetic screening amongst at-risk relatives (Burns et al., 2018; Ingles, Bagnall, et al., 2018; Ingles & Semsarian, 2018).

Many factors have been identified which influence family communication about genetic risk. These include complicated family dynamics, guilt, anxiety and gender (Barsevick et al., 2008; Burns et al., 2016; Christiaans et al., 2008; Claes et al., 2003). In addition, the literature and clinical experience highlights loss of contact with relatives and geographically distant relatives is a commonly cited and significant issue (Healey et al., 2017; Young et al., 2017). Importantly, much of this literature comes from the inherited cancer context. Inherited heart disease has the unique risk of sudden cardiac death, which should be considered when discussing communication of inheritance risk. Initial discussions surrounding a diagnosis of an inherited heart disease are often focused on clinical management of the proband themselves but should highlight the importance of family screening adherence (Burns et al., 2018; Hudson et al., 2018).

Studies focused on the inherited heart disease patient population aimed at addressing family communication show varying results. One recent study amongst HCM probands found 80% of first-degree relatives were informed of their genetic risk with probands acknowledging the unique process of communication for each family but perceiving disclosure of risk information as *‘imperative’ (Hudson et al*., *2018)*. In addition, we have conducted a study in long QT syndrome (LQTS) patients which demonstrated 10% of probands had not disclosed relevant risk information to at least one first-degree relative (Burns et al., 2016).

Evaluating genetic counseling interventions, particularly related to family communication is difficult. This may account for some of the ambiguity around the best practice approach. There is little agreement about suitable outcome measures to assess the effectiveness of a particular genetic counseling intervention (McAllister et al., 2011; Payne et al., 2008). Here we aimed to address the issue commonly referred to as passive non-disclosure, whereby relatives intend to disclose and communicate relevant information and do not actively choose non-disclosure. In spite of this however, communication still does not occur (Gaff et al., 2005). One contributing factor may be the information provided to probands and their knowledge of the appropriate information. Therefore, we aimed for our communication aid to improve knowledge and the information provided regarding family screening. In spite of the time spent with probands during the study, positive satisfaction and outcome scores, good confidence with genetic knowledge alongside a reasonable mean genetic knowledge score (60%), up to 17-29% of first-degree relatives amongst this cohort remain uninformed of their risk.

Essentially all outcomes in the study were non-significant. Design of our intervention may have addressed the wrong aspect of family communication about genetics. As discussed, family communication about genetics is complex and multifaceted, and in choosing to target ability and confidence as important contributors to family communication we may have missed a more appropriate outcome. Interpretation of these results and the literature, highlights the complexity, intricacies and personal and family dynamics that may play a significant role beyond knowledge in the process of family communication. A more tailored approach, addressing individual family needs and drawing upon more tools for supporting communication among families is likely needed.

Nonetheless, overall satisfaction and outcome scores were good. In addition, the cardiac genetic counselors using the communication aid found the aid to be clinically useful and commented that it facilitated the communication of genetic results to probands. In fact, it was identified that returning genetic results without use of the communication aid felt it was lacking after commencement of this study. Though data were not collected systematically, questions raised by the patients during the genetic counseling sessions with the communication aid reflected both a positive experience and firm grasp of the information provided for probands themselves.

## CONCLUSION

We highlight that in spite of satisfaction with services and the information provided, family communication was not improved by the genetic counselor-led intervention. Complex family dynamics, interpersonal family relationships and the proband’s own beliefs about whom they should communicate with all contribute to family communication about genetics. Interventions to support family communication and ensure all at-risk relatives are appropriately informed will require multifaceted approaches, allowing a tailored offering of support and tools to families.

## Data Availability

All data produced in the present study are available upon reasonable request to the authors and in accordance with ethical approvals.

## DATA AVAILABILITY

Additional information and data from this manuscript is available on reasonable request.

## AUTHOR CONTRIBUTIONS

*Authors CB and JI confirm that they had full access to all the data in the study and take responsibility for the integrity of the data and the accuracy of the data analysis*. *All of the authors gave final approval of this version to be published and agree to be accountable for all aspects of the work in ensuring that questions related to the accuracy or integrity of any part of the work are appropriately investigated and resolved*.

## ETHICS DECLARATION

Informed consent was provided by participants of this study. This study was approved by the Ethics review Committee (RPAH Zone) of the Sydney Local Health District (protocol number X16-0030) and all procedures were followed in accordance with the Helsinki Declaration of 1975, as revised in 2000.

